# Venezuelan equine encephalitis complex, Madariaga and Eastern equine encephalitis viruses genome detection in human and mosquito populations

**DOI:** 10.1101/2022.04.04.22271864

**Authors:** Jean-Paul Carrera, Dimelza Araúz, Alejandra Rojas, Fátima Cardozo, Victoria Stittleburg, Ingra Morales Claro, Josefrancisco Galue, Carlos Lezcano-Coba, Filipe Romero Rebello Moreira, Luis Felipe-Rivera, Maria Chen-Germán, Brechla Moreno, Zeuz Capitan-Barrios, Sandra López-Vérges, Juan Miguel Pascale, Ester C. Sabino, Anayansi Valderrama, Kathryn A. Hanley, Christl A. Donnelly, Nikos Vasilakis, Nuno R. Faria, Jesse J. Waggoner

## Abstract

Eastern equine encephalitis virus (EEEV), Madariaga virus (MADV) and Venezuelan equine encephalitis virus complex (VEEV) are New World mosquito-borne alphaviruses and cause severe neurological disease in human and equine hosts. However, their detection during the acute phase is complicated by non-specific clinical manifestations and lack of available diagnostic tools. To develop and clinically evaluate rRT-PCRs for VEEV complex, MADV and EEEV, primers and probes were designed from publicly available whole-genome sequences. The rRT-PCRs were validated using 15 retrospective serum samples from febrile patients collected during the 2015 and 2017 alphavirus outbreaks in Panama. In addition, the protocol was validated with 150 mosquito pools from 2015, and with 118 samples from prospective disease surveillance from 2021 and 2022. The rRT-PCRs detected VEEV complex RNA in 10 samples (66.7%) from the 2015 and 2017 outbreaks, and in one of these ten samples, both VEEV complex and MADV RNAs were detected. Additionally, VEEV complex RNA was detected in 5 suspected dengue from prospective disease surveillance. The rRT-PCR assays detected VEEV complex RNA in 3 from *Culex* (*Melanoconion*) *vomerifer* pools, 2 of which yielded VEEV isolates. Untargeted sequencing and phylogenetic analysis identified VEEV ID subtype in seven VEEV complex RNA positive sample. The VEEV complex, MADV and EEEV rRT-PCRs provide accurate detection while yielding significant benefits over currently available molecular methods. Our results suggest that 11.9% of suspected dengue cases in Panama are VEEV infections.

## Introduction

New World alphaviruses (*Togaviridae*, genus *Alphavirus*) comprise a diverse group of mosquito-borne viruses that can cause severe disease in humans, including the Venezuelan equine encephalitis virus complex (VEEV complex), Madariaga virus (MADV), and eastern equine encephalitis virus (EEEV)(1). These have single-stranded 11.7kb RNA genomes that encode four non-structural proteins (nsP1 to nsP4) and five structural proteins (capsid, E1, E2, E3 and 6K) (1, 2). VEEV complex, MADV and EEEV persist in sylvatic-enzootic cycles throughout the Americas and are transmitted to humans by *Aedes spp*., *Psorophora spp*. and *Culex spp*. mosquitoes (2, 3).

Serologic and molecular evidence of VEEV complex infections have been identified throughout tropical regions of Central and South America, suggesting VEEV complex infections may be relatively common but remain largely underdiagnosed(2). There is a high genetic variability within the VEEV complex, with at least 14 different viral subtypes identified to date (2). VEEV complex subtypes have been associated with large epizootic outbreaks involving equids and humans (VEEV subtypes IAB and IC)(1, 2). Although most infections in humans are asymptomatic or subclinical, patients may develop acute febrile illness with headache, myalgias, arthralgias, nausea, and vomiting (4, 5), which may progress to severe disease including encephalitis and the development of long-term neurologic sequelae(5, 6).

MADV, previously classified as South American EEEV, is an emerging virus that was first associated with large outbreaks in 2010 in the Darién province of Panamá (5), where VEEV subtype ID has also been detected(7). Before the 2010 outbreak in Panama, MADV was mostly associated with equine disease and only a few human infections had been detected in Trinidad and Tobago and Brazil(8, 9). This contrasts with the epidemiological profile of the North American EEEV, which is associated with severe and fatal human cases(3). Diagnostic tools are not widely available for MADV, and its prevalence outside of the Darién province in Panama remains poorly characterized(10). Geographic expansion of the Panamanian MADV strain into Northeast Brazil and Haiti has been reported recently, highlighting the potential of MADV to invade new areas (11, 12).

Accurate detection of VEEV complex, MADV and EEEV infections in the acute phase is complicated by their non-specific clinical manifestations and lack of widely available diagnostic tools. Antigen-based methods are currently unavailable, and although serology can confirm the diagnosis, this requires paired acute and convalescent samples (1, 5). Current molecular tests lack optimal performance characteristics necessary for routine diagnostic testing (13–19). Molecular assay designs have been complicated by the genetic variability of the VEEV complex viruses(2). VEEV complex infections are often misdiagnosed as dengue virus, due to overlapping clinical symptoms during the acute phase (2), and the recent classification of MADV as a separate virus species and human pathogen further complicates the clinical diagnosis of both(3). Common molecular tests for the VEEV complex employ pan-alphavirus primers to amplify a region of 400-500 nucleotides region of the genome with subsequent identification of the viral complex or species by sequencing or nested polymerase chain reaction (PCR)(5, 13, 14, 16, 18–20). Such methods are laborious and increase opportunities for laboratory contamination. In addition, pan-alphavirus primers and conventional reverse transcription-polymerase chain reaction (RT-PCR) chemistry may be less sensitive than real-time RT-PCR (rRT-PCR). Moreover, few rRT-PCR methods have been reported in the literature to date to detect and differentiate VEEV complex and MADV (21).

The primary objective of the current study was to design sensitive rRT-PCRs diagnosis assay for the VEEV complex and MADV. As a secondary objective, a duplex MADV/EEEV rRT-PCR was developed to differentiate these two pathogens. The two assays, for VEEV complex and MADV/EEEV detection, were evaluated in a set of clinical samples from an alphavirus outbreak in Panamá and prospective disease surveillance. Finally, we characterized viral species, subtype and genotype from selected rRT-PCR, positive samples from humans and mosquitoes collected during the 2015 and 2022 alphavirus outbreaks in Panama using metagenomic sequencing approach.

## Materials and Methods

### Ethics statement

The use of human samples used for protocol validation was approved by the Panamanian Ministry of Health (protocol number 2077), Gorgas’s Institutional Review Board (IRB) (protocol: 335/CBI/ICGES/21), Emory University IRB (IRB00097089), and the Ethics Committee of the Instituto de Investigaciones en Ciencias de la Salud, Universidad Nacional de Asunción (P06/2017). Prospective disease surveillance was approved by the Gorgas’s IRB (protocol:073/CBI/ICGES/21).

### Data availability

All the data used for human and mosquito validation are contained within the manuscript. Accessions numbers of newly derived genomes are XXX. Accessions numbers and strains information of sequences used for primer design are shown in Supplementary Appendix.

### VEEV complex, EEEV and MADV rRT-PCR design

Separate alignments were prepared for the VEEV complex, EEEV and MADV using all publicly available complete genome sequences from the NCBI GenBank (22) and aligned with MegAlign software (DNASTAR, Madison, WI). The VEEV complex alignment included all complete genomes from the following viruses: Cabassou, Everglades, Mosso das Pedras, Mucambo, Pixuna, Rio Negro, Tonate, and VEEV subtypes (IAB, IC, ID, and IE). The VEEV complex alignments were compiled in 2016 (n=121 sequences); a similar alignment for MADV was compiled in 2019 (n=32). Primers and probes were designed with Primer3 software (primer3.ut.ee) such that each oligonucleotide contained ≤1 degenerate base and matched ≥ 95% of available sequences for a given virus. In silico primer/probe specificity was checked by aligning sequences in BLAST (blast.ncbi.nlm.nih.gov) against (i) all available sequences and (ii) only alphavirus sequences while excluding the VEEV complex or MADV, respectively. Due to the similarity between MADV primers and EEEV sequences, all available EEEV complete genome sequences (n=441) were aligned and separate MADV and EEEV probes were designed for an rRT-PCR duplex assay. Alignments for each virus were repeated with all sequences available in September 2021 to confirm primer and probe sequences in contemporary strains.

### rRT-PCR assay performance and optimization

rRT-PCRs were performed in 25µL reactions using the SuperScript III Platinum One-Step Quantitative RT-PCR Kit (Thermo Fisher, Waltham, MA) with 5µL of the nucleic acid template. The analytical evaluation was performed on a Rotor-Gene Q instrument (Qiagen, Germantown, MD), and the validation with serum and mosquito pool samples was performed on an ABI7500 (Thermo Fisher). Cycling conditions were consistent with previous laboratory protocols: 52 °C × 15 min, 94 °C × 2 min, and 45 cycles of 94 °C × 15 s, 55 °C × 40 s (acquired in all channels), and 68 °C × 20 s (1-4). Primer and probe sets were evaluated in singleplex reactions containing 200nM of each oligonucleotide and genomic RNA or quantified ssDNA containing the target region. Primer/probe sets were selected to generate the most sensitive detection based on cycle threshold (Ct) values, with preserved specificity. Primer and probe concentrations in the final reaction were then adjusted between 100nM and 400nM to optimize assay sensitivity. Primers were obtained from Integrated DNA Technologies (IDT, Coralville, Iowa); probes were obtained from Biosearch Technologies (Hoddesdon, United Kingdom). VEEV subtype IC and EEEV genomic RNAs were purchased from Vircell Microbiologists (Granada, Spain). Quantified ssDNA containing the assay target region (IDT, Coralville, Iowa) was used for all viruses to quantify analytical performance. For ssDNA synthesis, target region sequences were selected from specific strains of VEE subtype IAB (Accession number KC344505.2) and subtype IV (Pixuna virus, Accession number NC_038673.1), MADV (Accession numbers MH359233.1 and KJ469626.1), and EEEV (Accession number KX029319.1).

### Analytical evaluation

The linear range of the assay was determined by testing synthesized targets from each reference strain in quadruplicate at 8.0, 6.0, 4.0, 2.0, and 1.0 log_10_ copies/μL. The linear range was defined as the range of concentrations at which 1) all replicates were detected and 2) the linear regression of Ct values (y, dependent variable) versus log_10_ copies/ μL (x, independent variable) generated an R^2^ ≥0.99. For the VEEV complex, the lower limit 95% detection (95% LLOD) was determined by testing 10 replicates of 2-fold serial dilutions from 200 to 25 copies/μl. For MADV and EEEV, 95% LLOD was calculated directly from the linear range. Assay exclusivity was evaluated by testing genomic RNA from the following viruses (strain in parentheses, if designated): Rift Valley fever (h85/09); Zika (ZIJV; MR766); dengue virus serotype 1 (DENV1, Hawwai 1944), DENV2 (NGC), DENV3 (Sleman/78), and DENV4 (H241); chikungunya virus (CHIKVR80422); Mayaro virus (MAYV; ARV 0565, INHRR 11a-10); yellow fever virus (YFV; 17D and Asibi strains); West Nile virus (WNV; NAL); St. Louis encephalitis virus (SLEV; GML 902612, CorAn 9275); tick-borne encephalitis virus (TBEV; Japanese encephalitis virus (JEV); Semliki Forest virus (SFV); Ross River virus (RRV); Getah virus (GETV); Barmah Forest virus (BFV); and Una virus (UNAV) (5). Specificity was also evaluated by testing 56 serum samples from locations without known transmission of VEEV or MADV. These included 8 samples collected from patients in Georgia, USA, without known travel history, and 48 samples from individuals with an acute febrile illness in Asunción, Paraguay. The latter samples have been described in detail elsewhere (23). Total nucleic acids were extracted from 200µL of serum on an EMAG instrument (BioMérieux, Durham, NC), eluted in 50µL and run in the VEEV complex and MADV/EEEV rRT-PCRs.

### Statistics

Linear regression fitting and linear range calculations, including R^2^ of the best-fit line were performed in Excel (Microsoft, Redmond, WA). Concentrations and replicates in the linear range study that did not yield Ct values were not included in the linear regression analysis. Probit analyses were performed using MedCalc, v20.013 (MedCalc Software, Belgium).

### Protocol validation with acute human samples

Acute human samples used in the protocol validation were collected in communities of Darien, the eastern most province in Panama, during three alphavirus outbreaks in 2015, 2017. Cases identified in 2015 and 2017 were detected in the communities of Metetí, Cemaco, Tucutí, Yaviza, Nicanor, La Palma and El Real de Santa María (Figure 1A). The Darien province borders Colombia and encompasses the Darien Gap, and the Darien National Park, a UNESCO-designated World Heritage Site(24).

**Figure 1.**
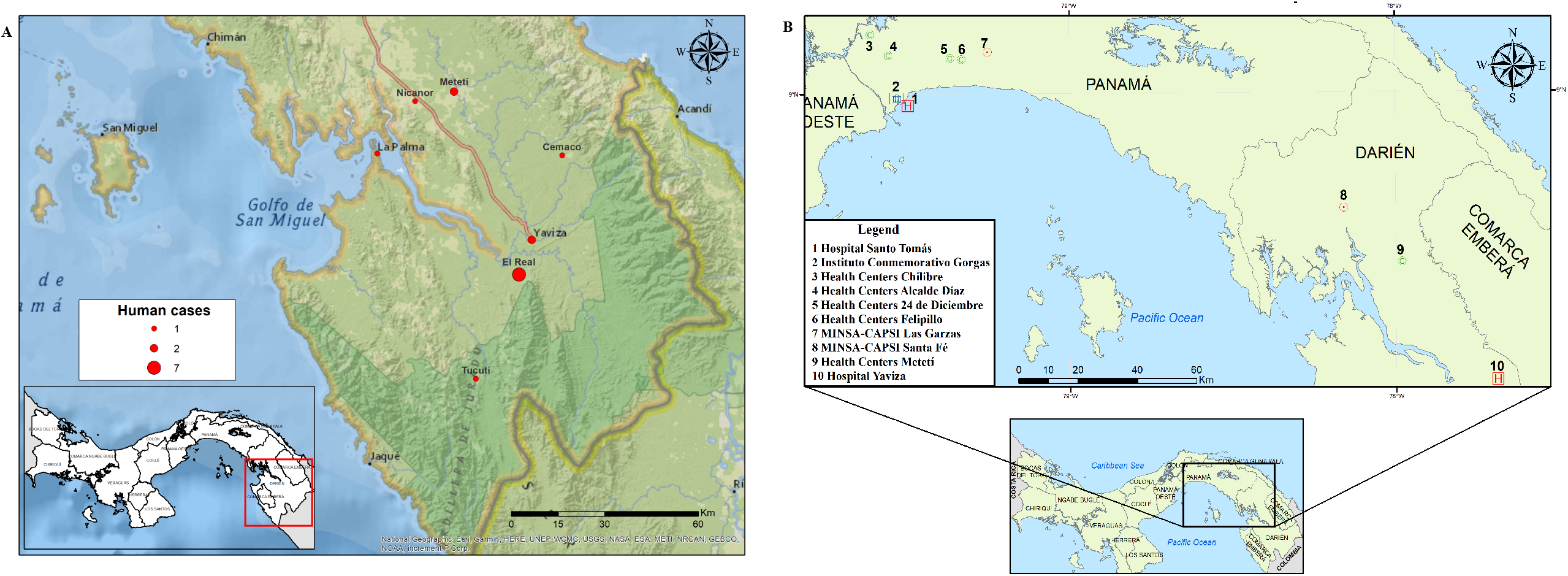
Map with the distribution of VEEV human cases in Darien province in 2015 and 2017 and Health centres in Panama and Darien Provinces. **A**. Distribution of VEEV cases used for protocol validation. Red dots represent the number of cases reported by locality. B. Distribution of Health centers used for prospective febrile surveillance in Panama and Darien provinces. Map was created with ArcGIS Desktop 10.6 using shapefiles from Esri. Data sources for the shapefiles include Esri, Garmin International Inc., US Central Intelligence Agency, and National Geographic Society (39).

### Patient recruitment in 2015 and 2017

Febrile patients were identified during an enhanced surveillance program by our outbreak response team using house-by-house visits during the 2015 and 2017 outbreaks. Blood samples were drawn from patients that met the case definition during the outbreak investigation. The case definition of a suspected case included fever and headache, while a probable case was defined as a suspected case plus somnolence, lethargy, or convulsions. Blood samples were centrifuged in the field, and serum was stored in liquid nitrogen for transportation to the Gorgas Memorial Institute of Health Studies in Panama City.

### Prospective acute disease surveillance in 2021 and 2022

In 2021 an effort for surveillance of emerging pathogens was established in Panama as part of the USA-National Institute of Allergy and Infectious Diseases (NIAID), Centers for Research in Emerging Infectious Diseases Network initiative. The **C**oordinating **R**esearch on **E**merging **A**rboviral **T**hreats **E**ncompassing the **N**eotropics (CREATE-NEO) in Panama undertakes acute febrile surveillance across ten Health Centers in Panama and Darien Provinces (Figure 1 B) (https://www.utmb.edu/createneo/home/create-neo-home). Cases, with malaria, human immunodeficiency virus (HIV), hepatitis B (HBV) and hepatitis (HCV), and >5 and <75 years old, presenting with no more than 7 days with rash, and at least one of the following symptoms: fever, myalgia, arthralgia, periarticular edema, and conjunctivitis were recruited, evaluated and interviewed, to obtain clinical, and demographics characteristics and ethic consent at each health center.

### Laboratory testing for acute disease surveillance

Acute samples (0-5 days) were first screened against DENV, CHIKV and ZIKV virus using rRT-PCR as described previously (25), followed by testing with the newly designed MADV/VEEV rRT-PCR.

### Mosquito collection

Mosquito collections were performed in a forested section of 100 × 100 meters within El Real de Santa María (Figure 1 A). Acute VEEV cases were identified during the 2015 outbreak response in El Real de Santa María. Mosquito collections were undertaken with CDC light traps and performed over a period of 12 hours, from 6:00 pm to 6:00 am at a height of 1.5 m above ground level. Traps (BioQuip Products, Rancho Dominguez, CA) were baited with octanol and CO_2_ for the encephalitis vector survey. The following day, mosquitoes were collected in the field and transported to the base camp where they were anesthetized, identified to the species level using taxonomic keys (26), and placed in cryovials for storage in liquid nitrogen. Mosquitos were grouped at the species level and a maximum of 20 individuals were used to produce mosquito pools for rRT-PCR, serology, and isolation.

### Alphavirus serology of clinical samples

All human serum samples were tested in duplicate for IgM antibodies to MADV and VEEV antigen using an enzyme-linked immunosorbent assay (ELISA) and confirmed by a plaque-reduction neutralization test (PRNT). For the ELISA, sucrose-acetone antigens were prepared from MADV- (prepared by Dr. Robert Shope at the Yale Arbovirus Research Unit in August 1989) and VEEV- (strain TC-83) infected mouse brain. For the PRNT, we used chimeric Sindbis virus SINV/MADV (derived from Brazilian MADV strain BeAn436087 and shown to be an accurate surrogate for MADV in these assays (27) and TC83, an attenuated vaccine strain of VEEV closely related to subtype ID strains that circulate in Panama (7). The neutralizing antibody titer was determined as the reciprocal of the highest dilution that reduced plaque count by 80% (PRNT_80_).

### Viral isolation from mosquito pools

Mosquito pool homogenates were prepared with 20 – 50 mosquitoes in 2 mL of minimum essential medium supplemented with penicillin and streptomycin, and 20% fetal bovine serum, homogenized using a Tissue Lyser (Qiagen, Hidden, Germany), and centrifuged at 12000 rpm for 10 mins. A total of 200 μL of serum or mosquito homogenate was inoculated in each of two 12.5-cm^2^ flasks of Vero cells. Samples were passed twice and monitored for cytopathic effect (CPE).

### Generic alphavirus RT-PCR for human and mosquito samples

Viral RNA was extracted from human sera and mosquito pool homogenates using QIAamp RNA viral extraction kit (Qiagen, Valencia, CA). Viral RNA from mosquitoes was also extracted using the Macherey-Nagel extraction kit (Düren, Germany). Sera and mosquito homogenates were tested in 25µL reactions for alphaviruses using a universal alphavirus RT-PCR, as previously described (19).

### Viral metagenomic sequencing

To confirm virus species, subtype and genotype, we sequenced seven selected VEEV complex rRT-PCR positive mosquito and human samples from 2015 and 2022 using SMART-9N metagenomic sequencing ad previously described (28). In brief, viral RNA was treated with TURBO DNase (Thermo Fisher Scientific, USA) and concentrated with Zymo RNA clean & concentrator-5 (Zymo Research, USA) following protocol instructions. cDNA synthesis and PCR was performed as described (28). Fifty ng of the quantified PCR products were pooled using EXP-NBD104 (1-12) and EXP-NBD114 (13-24) Native Barcoding Kits (ONT, UK). Sequencing libraries were generated using the SQK-LSK109 kit (ONT, UK) and loaded onto FLO-MIN106 flow cells on the GridION device (ONT, UK). Sequencing base-calling and demultiplexing was performed by MinKNOW with the standard 48-hour run script (ONT, UK). Demultiplexed FASTQ files were aligned and mapped to the VEEV reference genome (GenBank accession no. NC_001449.1) using minimap2 version 2.28. (29) and converted to sorted BAM file using SaMtool(30). NanoStat version 1.1.2.4,(31) and Tablet (32) were used to compute genomic statistics. Variants were detected with medaka_variants and consensus sequences were built with marging_medaka_consensus (ONT, UK). Genomic regions with <20x coverage were masked.

### VEEV Phylogenetic analysis

All available VEEV genome sequences, in GenBank, representing all antigenic complex were selected to construct the alignment, duplicated sequences, partial sequences and overlapping sequences were removed. Finally, the novel complete or near complete VEEV genome sequences (n=7) were aligned with 132 representative VEEV genomes retrieved from NCBI GenBank using MAFFT version 7 (33). Selection of the best-fitting nucleotide substitution model and maximum likelihood phylogenetic reconstruction were performed with IQ-Tree v2.2.0.3(34). Statistical robustness of the tree topology was assessed with 1,000 ultrafast bootstrap replicates.

## Results

### rRT-PCR analytical evaluation

Primers and probes for the VEE complex singleplex and MADV/EEEV duplex rRT-PCRs are shown in Table 1 along with the optimized final reaction concentrations. The linear range for each assay extended from 2.0 to 8.0 log_10_ copies/µL (Figure 2A-D). For the VEEV complex assay, the linear range was evaluated with ssDNA for subtypes IAB and IV and RNA from subtype IC (2.0 to 5.0 log_10_ copies/µL; Figure 1A-B). The 95% LLODs, expressed in copies/µL, were: VEEV subtype IAB, 120; VEE subtype IV, 110; MADV, 19; EEEV, 19. Assay exclusivity was evaluated by testing genomic RNA from VEEV subtype IC, EEEV, and a set of arboviruses, including flavi-, bunya-, and alphaviruses on a single run of the VEEV complex and MADV/EEEV rRT-PCRs. VEEV complex and EEEV only yielded signals in the respective assays for these viruses. None of the other tested viruses generated signal in either assay. In addition, none of the 56 serum samples from Georgia, USA, or Asunción, Paraguay, tested positive in either assay.

**Table 1.**
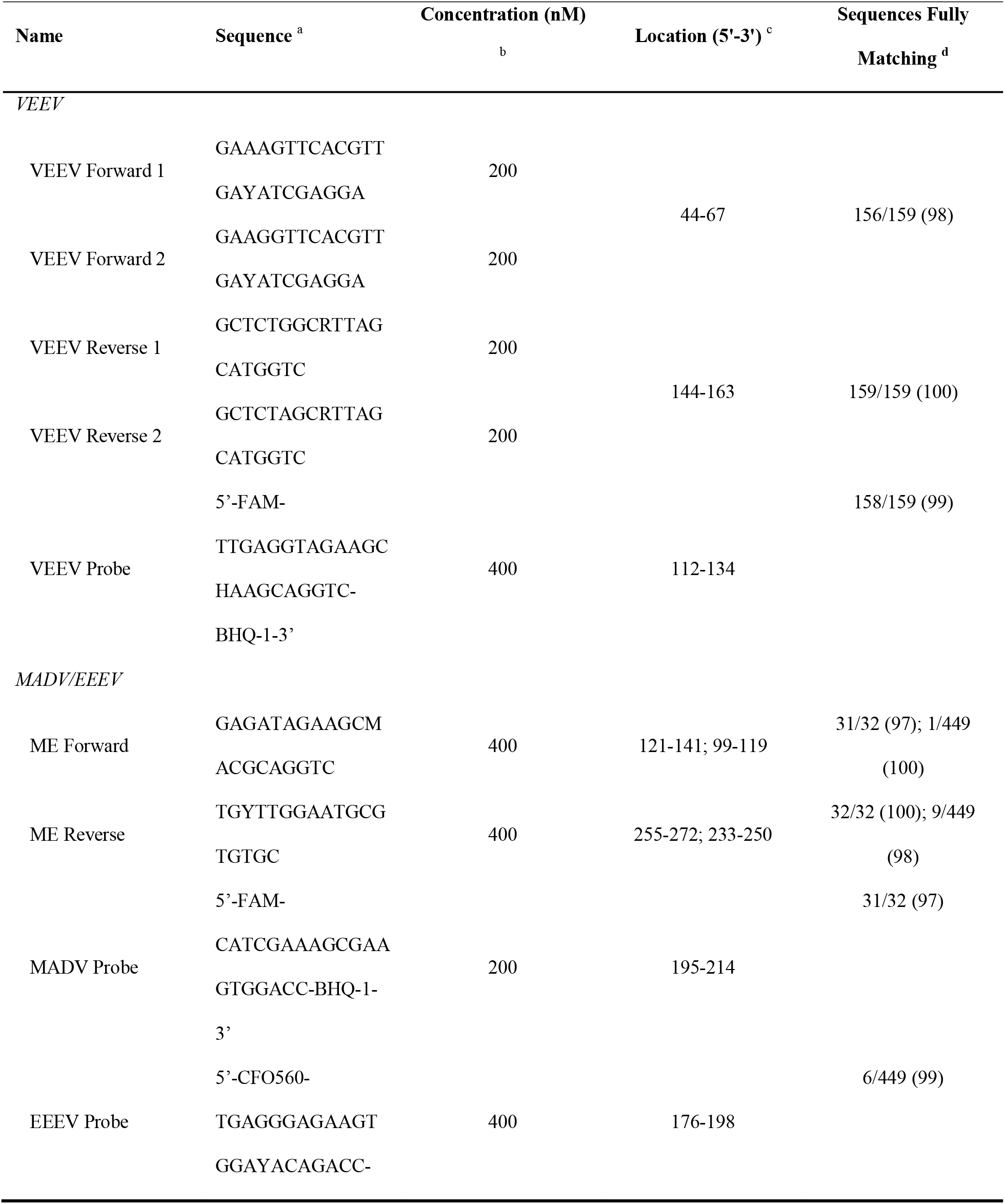
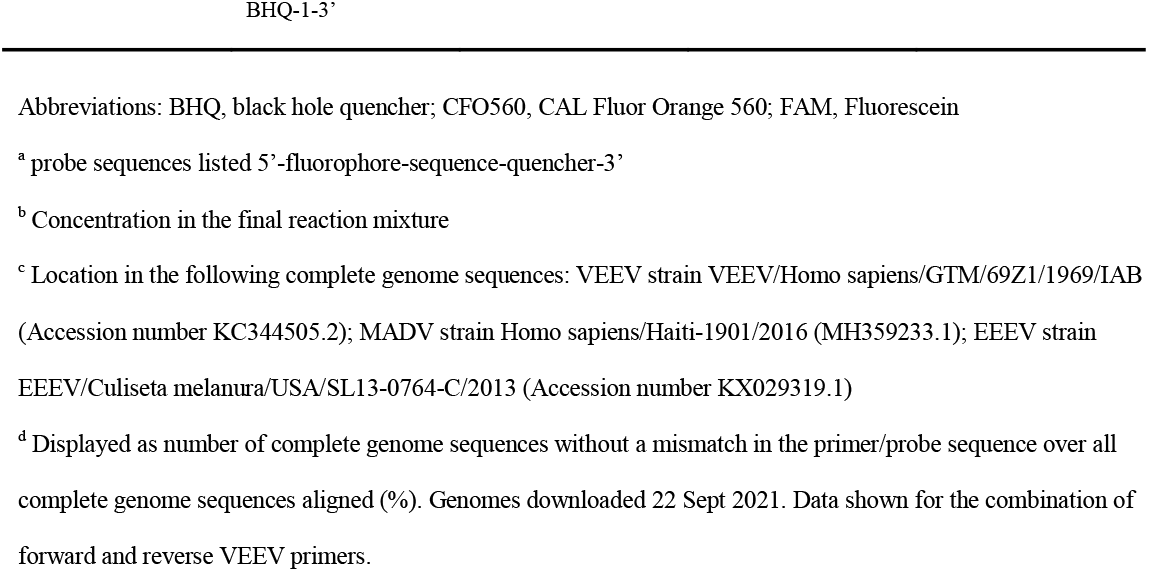
Primers and probes in the VEEV and MADV/EEEV rRT-PCRs.

**Figure 2.**
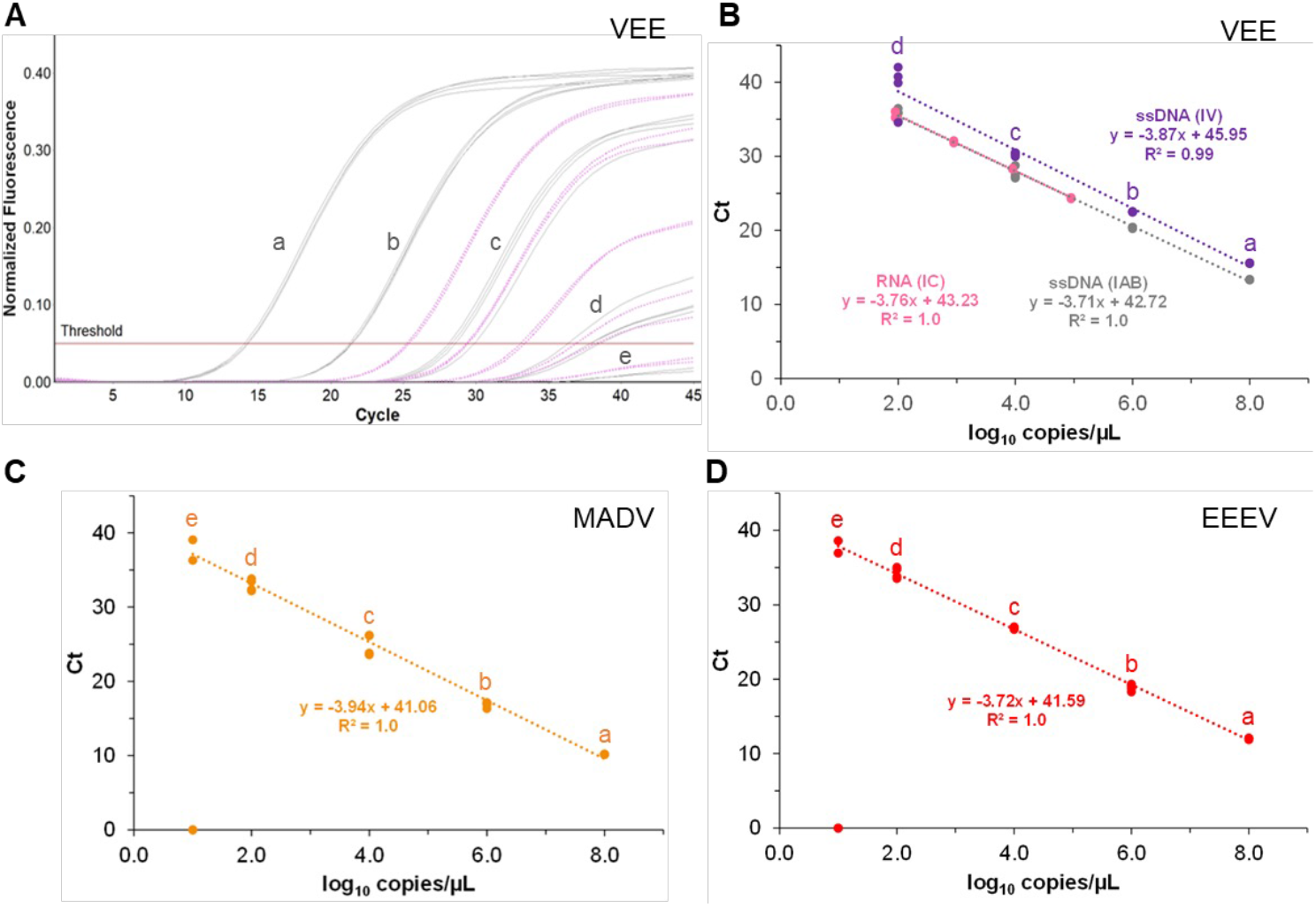
VEE and MADV/EEEV rRT-PCR linearity and limit of detection. **A**) Amplification curves across the linear range of the VEE complex rRT-PCR with ssDNA (gray curves, subtype IAB) and RNA (pink dotted curves, subtype IC). **B**) Linearity and limit of detection for VEE were evaluated with ssDNA from subtypes IAB (grey) and IV (purple) and RNA from subtype IC (pink). (**C-D**) Linearity and limit of detection for MADV © and EEEV (D) were evaluated using ssDNA. For each virus, ssDNA was tested in quadruplicate at 8.0, 6.0, 4.0, 2.0 and 1.0 log_10_ copies/µL (labelled a-e, respectively). 10-fold dilutions of VEEV subtype IC RNA were tested in duplicate starting at the highest concentration available (5.0 log_10_ copies/µL). All data points are displayed, including 2/4 MADV and EEEV replicates that did not yield cycle threshold (Ct) values and were excluded from the linear regressions.

### Validation with clinical samples

A total of 15 febrile patients from 2015 and 2017 alphavirus outbreaks that met the suspected or probable case definition were used to validate the new molecular assays. Previously, a total of eleven (11/15) acute sera samples collected during the in 2015 and 2017 alphavirus outbreaks had tested positive using a generic alphavirus RT-PCR and were confirmed later by sequencing as VEEV-ID infections (17). In 2021, a second round of generic alphavirus RT-PCR using the same set of primers was run on these 15 stored samples, and all of them tested negative. Notably, using the newly designed rRT-PCR, we were able to detect 10 VEEV complex RNA positive samples (Ct range: 27 – 38), including two samples that had tested negative at the initial screening in 2017 (Table 2). Three of the VEEV complex rRT-PCR-positive samples were also anti-VEEV IgG and IgM positive, with only 0, 2, and 3 days since the onset of symptoms, respectively (Table 2). One sample was rRT-PCR positive for both VEEV and MADV viruses.

**Table 2.**
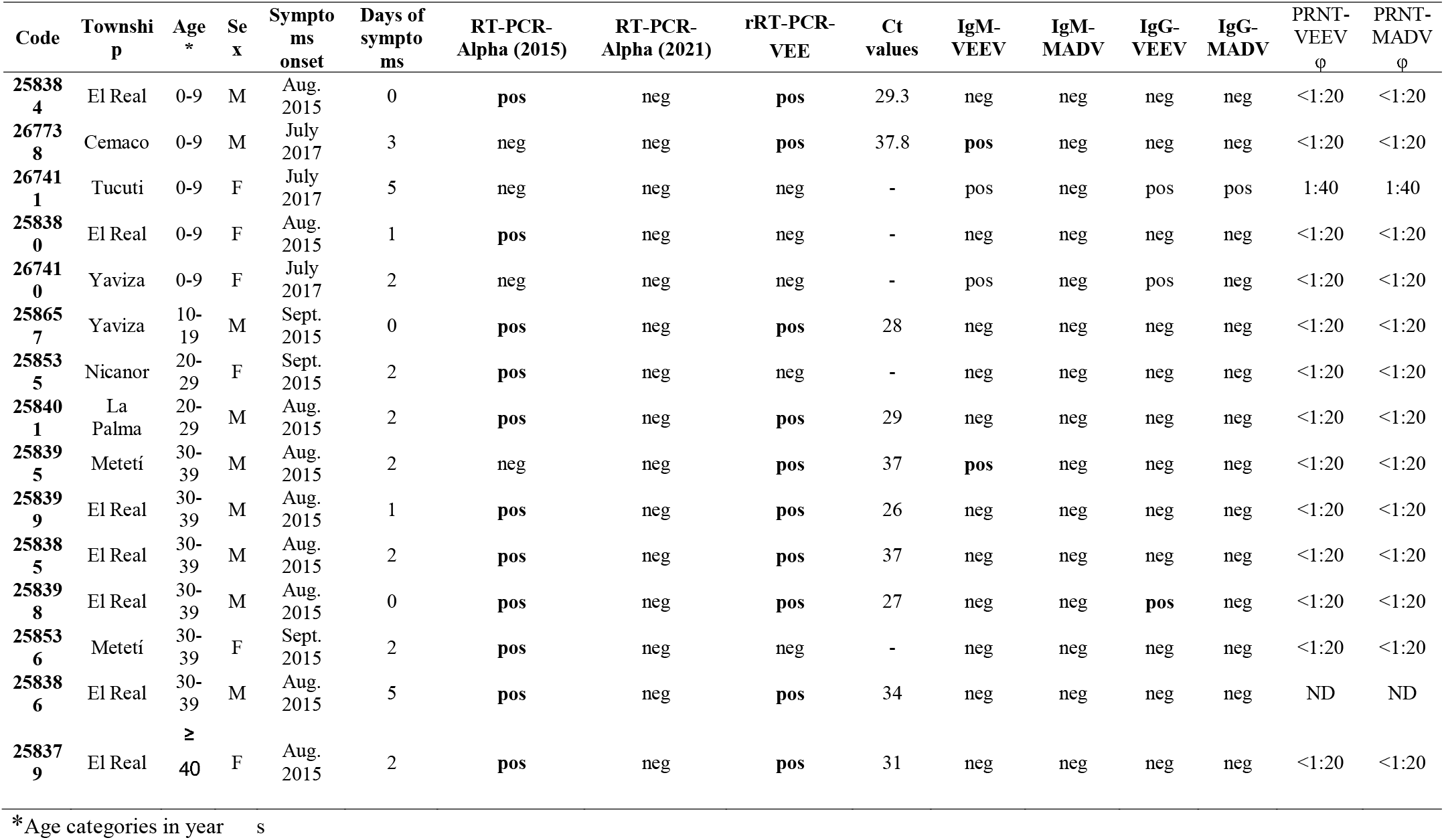
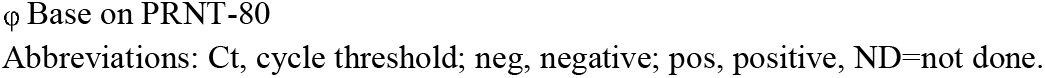
Characteristics and laboratory results of samples used for protocol validation patients and clinical samples and laboratory results. Acute samples selected from the 2015 and 2017 alphavirus outbreaks in Darien Province.

| Code | Township | Age * | Sex | Symptoms onset | Days of symptoms | RT-PCR-Alpha (2015) | RT-PCR-Alpha (2021) | rRT-PCR-VEE | Ct values | IgM-VEEV | IgM-MADV | IgG-VEEV | IgG-MADV | PRNT-VEEV $\phi$ | PRNT-MADV $\phi$ |
| --- | --- | --- | --- | --- | --- | --- | --- | --- | --- | --- | --- | --- | --- | --- | --- |
| 258384 | El Real | 0-9 | M | Aug. 2015 | 0 | pos | neg | pos | 29.3 | neg | neg | neg | neg | <1:20 | <1:20 |
| 267738 | Cemaco | 0-9 | M | July 2017 | 3 | neg | neg | pos | 37.8 | pos | neg | neg | neg | <1:20 | <1:20 |
| 267411 | Tucuti | 0-9 | F | July 2017 | 5 | neg | neg | neg | - | pos | neg | pos | pos | 1:40 | 1:40 |
| 258380 | El Real | 0-9 | F | Aug. 2015 | 1 | pos | neg | neg | - | neg | neg | neg | neg | <1:20 | <1:20 |
| 267410 | Yaviza | 0-9 | F | July 2017 | 2 | neg | neg | neg | - | pos | neg | pos | neg | <1:20 | <1:20 |
| 258657 | Yaviza | 10-19 | M | Sept. 2015 | 0 | pos | neg | pos | 28 | neg | neg | neg | neg | <1:20 | <1:20 |
| 258535 | Nicanor | 20-29 | F | Sept. 2015 | 2 | pos | neg | neg | - | neg | neg | neg | neg | <1:20 | <1:20 |
| 258401 | La Palma | 20-29 | M | Aug. 2015 | 2 | pos | neg | pos | 29 | neg | neg | neg | neg | <1:20 | <1:20 |
| 258395 | Metetí | 30-39 | M | Aug. 2015 | 2 | neg | neg | pos | 37 | pos | neg | neg | neg | <1:20 | <1:20 |
| 258399 | El Real | 30-39 | M | Aug. 2015 | 1 | pos | neg | pos | 26 | neg | neg | neg | neg | <1:20 | <1:20 |
| 258385 | El Real | 30-39 | M | Aug. 2015 | 2 | pos | neg | pos | 37 | neg | neg | neg | neg | <1:20 | <1:20 |
| 258398 | El Real | 30-39 | M | Aug. 2015 | 0 | pos | neg | pos | 27 | neg | neg | pos | neg | <1:20 | <1:20 |
| 258536 | Metetí | 30-39 | F | Sept. 2015 | 2 | pos | neg | neg | - | neg | neg | neg | neg | <1:20 | <1:20 |
| 258386 | El Real | 30-39 | M | Aug. 2015 | 5 | pos | neg | pos | 34 | neg | neg | neg | neg | ND | ND |
| 258379 | El Real | ≥ 40 | F | Aug. 2015 | 2 | pos | neg | pos | 31 | neg | neg | neg | neg | <1:20 | <1:20 |

\*Age categories in years
φ Base on PRNT-80
Abbreviations: Ct, cycle threshold; neg, negative; pos, positive, ND=not done.

### Prospective disease surveillance

A total of 118 febrile patients were recruited from November 16, 2021, to December 1, 2022. Of these 84 (71.2%) were acute patients with onset of symptoms ranging from 0-5 days. A total of 42 patients (50.0 %) were DENV1 positive. We detected VEEV RNA (Ct range: 15-20) in five patients (11.9%; 95% CI: 4.0 – 25.6) with suspected dengue infection, one of which was from a fatal case in 2022. Details and results of disease surveillance are presented in Figure 3.

**Figure 3.**
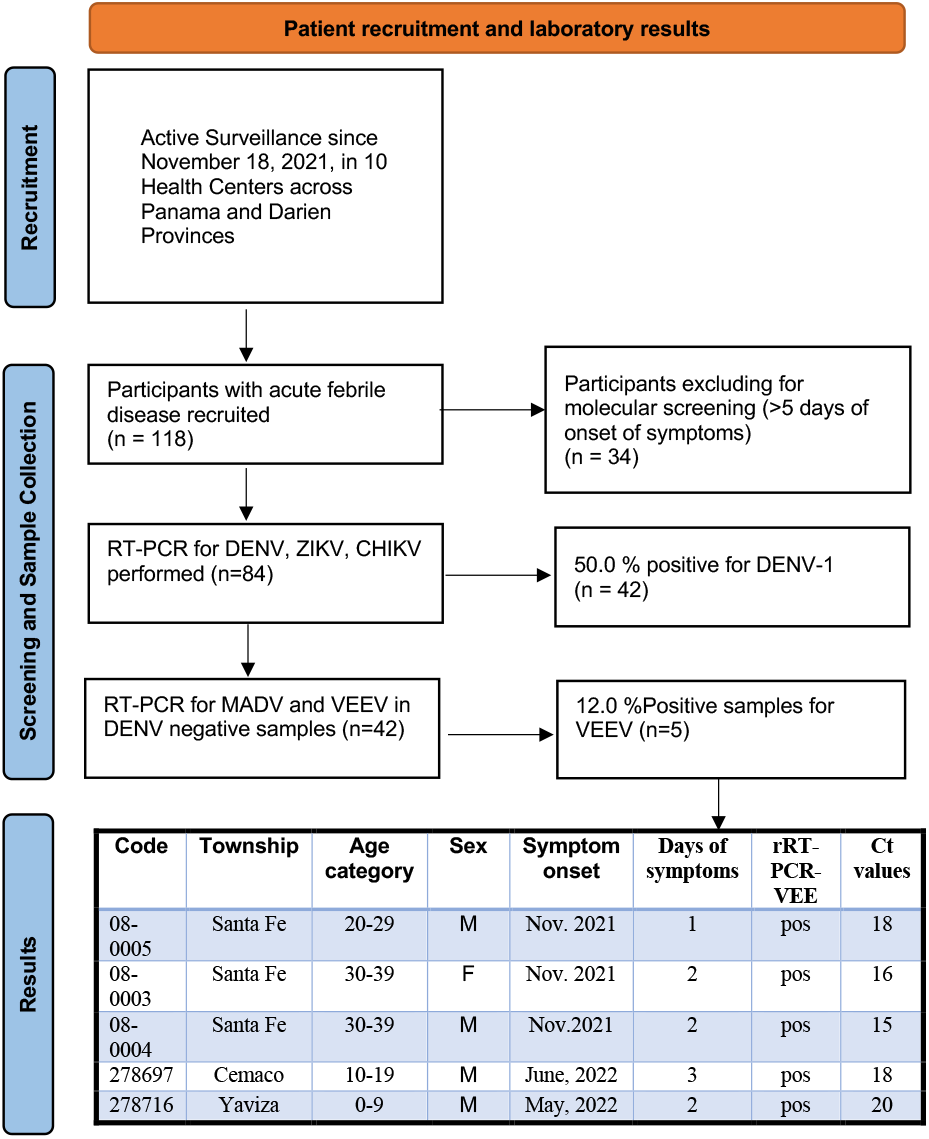
Flowchart of patient recruitment, characteristics and RT-PCR results of febrile patients detected throughout disease surveillance. Febrile patients were recruited from November 16, 2021, to December 1, 2022, in ten health care centers of Panama and Darien provinces.

### Viral detection in mosquito pools

A total of 1307 mosquitoes belonging to 35 species and 12 genera were collected in the community of El Real de Santa Maria, Panama, during a period of five days in 2015 (table 3). The most abundant mosquito species was *Coquilletidia* venezualensis (37.5%, n=490 of 1307) and *Culex Melanoconion vomerifer* (34,4%, n=450 of 1307). Mosquito species, number of individuals and pools are shown in Table 3. Of 150 mosquito pools, 3 *Cx. (Mel*.*) vomerifer* mosquito pools tested positive for VEEV by rRT-PCR (Ct range:26-30). Two of these rRT-PCR positive pools also yielded viral isolates.

**Table 3.** Mosquito species collected during the 2015 outbreak in El Real de Santa Maria, Panama.

| Mosquitos species | N | (%) | # Pools | VEE-rRT-PCR<br>Positive | MADV-rRT-PCR<br>Positive | Viral solates |
| --- | --- | --- | --- | --- | --- | --- |
| *Coquillettidia venezuelensis* | 490 | 37.5 | 29 | 0 | 0 | 0 |
| *Culex (Melanoconion) vomerifer* | 450 | 34.4 | 27 | 3 | 0 | 2 |
| *Culex (Melanoconion) pedroi* | 32 | 2.4 | 4 | 0 | 0 | 0 |
| *Aedes serratus* | 31 | 2.4 | 7 | 0 | 0 | 0 |
| *Aedes sp.* | 30 | 2.3 | 5 | 0 | 0 | 0 |
| *Culex (Melanoconion) sp.* | 30 | 2.3 | 6 | 0 | 0 | 0 |
| *Culex (Culex) interrogator* | 27 | 2.1 | 5 | 0 | 0 | 0 |
| *Anopheles trianulatus* | 23 | 1.8 | 2 | 0 | 0 | 0 |
| *Aedes eupoclamus* | 14 | 1.1 | 4 | 0 | 0 | 0 |
| *Culex (Culex) nigripalpus* | 14 | 1.1 | 3 | 0 | 0 | 0 |
| *Culex (Culex) sp.* | 14 | 1.0 | 4 | 0 | 0 | 0 |
| *Culex (Melanoconion) atratus* | 14 | 1.0 | 1 | 0 | 0 | 0 |
| *Culex (Melanoconion) adamesi* | 13 | 1.0 | 3 | 0 | 0 | 0 |
| Others* | 125 | 9.6 | 50 | 0 | 0 | 0 |
| Total | 1307 | 100 | 150 | 3 | 0 | 2 |

Species <1% abundance are listed as Others.

### VEEV Subtype identification

Three mosquito pools and 4 human samples (including one from a fatal case in 2022), that tested positive with the new VEEV complex rRT-PCR were sequenced using a virus untargeted approach (https://wellcomeopenresearch.org/articles/6-241). Twenty-fold genome coverage ranged from 45% to 100% (Table 4). Percentage of genome identity with VEEV reference strain ranged from 87.7% to 90.0% (Table 4), while identity with the Panamanian VEEV ID subtype prototype strain 3880 ranged from 96 to 97% (Table 4). Maximum likelihood phylogenetic analysis indicated that the new viral genomes cluster together with historical Panamanian VEEV ID subtype strains within the Panama/Peru genotype (bootstrap statistical support =100; Figure 4). Percentage of genome identity with VEEV reference strain ranged from 45% to 100% (Table 4).

**Table 4.** Metadata and sequencing statistics for selected VEEV complex RNA positive samples.

| ID | Collection date | Location | Host species | %genome coverage 20X | % Nt Identity with genome reference<sup>a</sup> | % Identity with strain 3880<sup>b</sup> |
| --- | --- | --- | --- | --- | --- | --- |
| 700677 | 2015 | Darien | *Culex (Mel.) vomerifer* | 100 | 89.8 | 92.1 |
| 700680 | 2015 | Darien | *Culex (Mel.) vomerifer* | 100 | 89.8 | 92.2 |
| 700732 | 2015 | Darien | *Culex (Mel.) vomerifer* | 100 | 90 | 92.3 |
| 258379 | 2015 | Darien | *Human* | 99.9 | 89.6 | 92 |
| 258398 | 2015 | Darien | *Human* | 70 | 88.7 | 90.7 |
| 258401 | 2015 | Darien | *Human* | 90.6 | 87.7 | 90 |
| 278716 | 2022 | Darien | *Human* | 45.98 | 88.1 | 90 |

<sup>a</sup>Genbank accession no. NC\_001449.1
<sup>b</sup>Panamanian VEEV ID subtype prototype strain 3880, GenBank accession no. L00930.1 Nt=Nucleotide.

**Figure 4.**
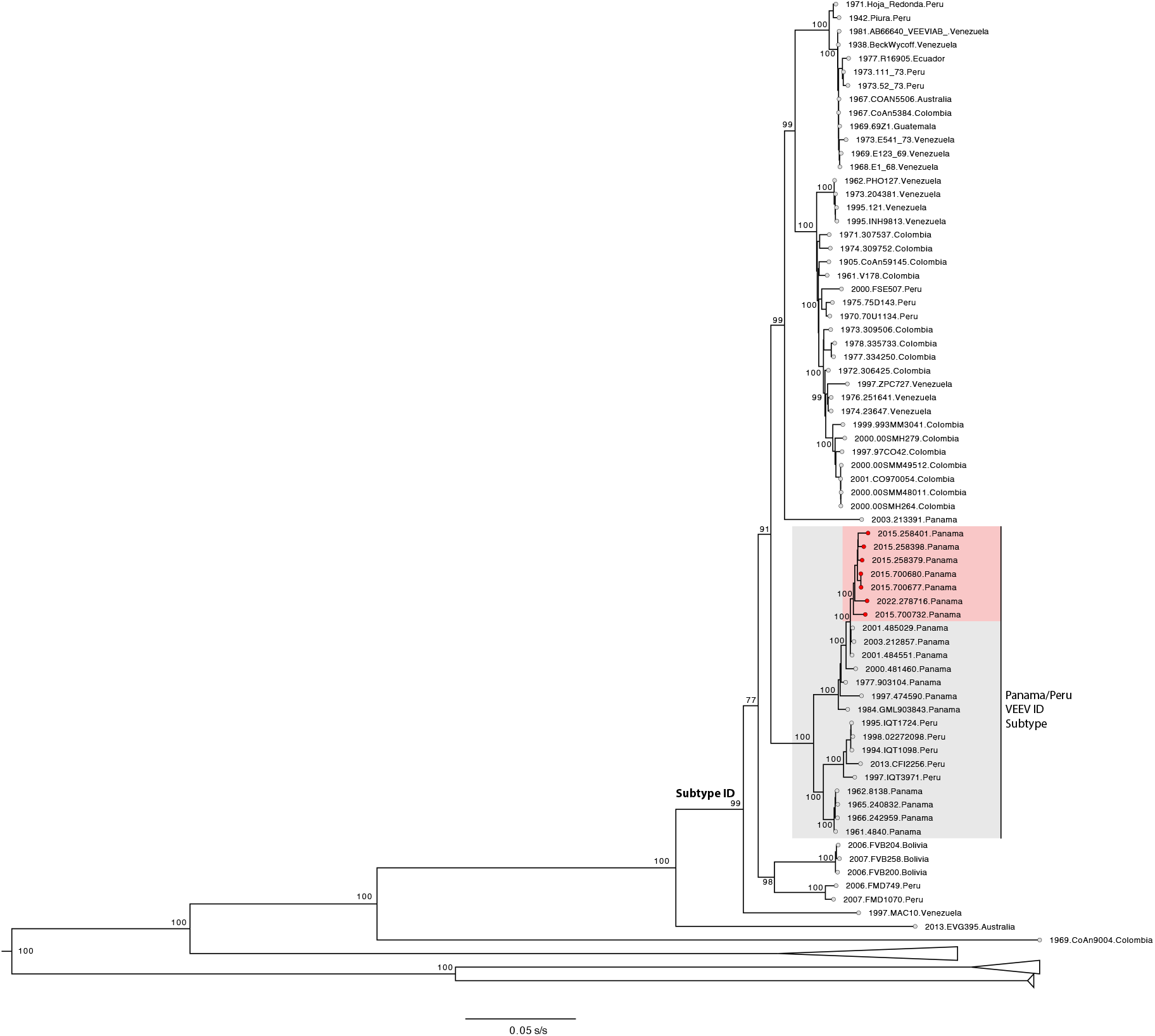
VEEV complex maximum likelihood phylogenetic tree. Maximum likelihood phylogenic was estimated using 139 complete or near complete VEEV genomes. Publicly available Panamanian VEEV ID subtype strains are highlighted in grey (n=) and genomes generated in this study (n=7) are highlighted in red. Bootstrap statistical support are shown for selected nodes. NCBI GenBank accessions numbers for the new VEEV genomes are: XX-XX.

## Discussion

The VEEV complex, MADV and EEEV viruses have been detected throughout the Americas and may account for a significant proportion of non-dengue acute febrile illness (2, 3, 5, 9). Although several assays have been developed for the molecular detection of VEEV subtypes and VEEV complex (14–20), many are laborious and time-consuming based in multiple PCR rounds or posterior genome sequencing that can be only implemented in laboratories with appropriate facilities (14–20). Co-circulation and the potential for co-infection with these viruses further complicates their detection, particularly when they present similar clinical presentations and there is a lack of available and convenient methods for detection of VEEV complex viruses and MADV (19). For example, VEEV subtype ID and MADV have both been identified in Panamá, and co-circulation of VEEV and MADV was detected in the Darién province of Panamá along the Colombian border (5–7). Typically, cases are often detected during the neurological phase of the disease (5, 35). At this stage, the virus has been cleared from the serum and diagnosis relies mostly on serological testing. Because alphaviruses can induce an IgM response that lasts up to 2 to 3 months, the detection of anti-VEEV or anti-MADV IgM alone could yield a misdiagnosis if seroconversion is not observed (5, 35).

We developed new accurate VEEV complex singleplex and MADV/EEEV duplex rRT-PCRs for the detection of viral RNA and real-time differentiation of clinical and mosquito samples. Using these assays, we detected VEEV ID subtype and MADV in samples that had tested negative with a reference RT-PCR (19). We were also able to identify an individual with a VEEV ID subtype - MADV co-infection, highlighting another advantage of our VEEV complex and MADV/EEEV rRT-PCRs over previous methods. Co-infection cases are epidemiologically significant and may have clinical relevance if associated with more severe disease (5). Our newly developed rRT-PCR assays can be rapidly incorporated into diagnostic algorithms in endemic regions. We show that the currently developed rRT-PCR allowed us to detect VEEV ID subtype RNA in samples collected from patients at least within the first 5 days of symptoms, whereas alphavirus IgM and IgG antibody responses usually develop >5–7 days post symptom onset (36). Interestingly, three patients with detectable VEEV complex RNA were also VEEV IgM and IgG-reactive, suggesting that VEEV re-infections are possible, a finding with potential implications for alphavirus vaccine development.

Our newly developed rRT-PCR allows the detection of the recently identified VEEV ID subtype in acute infections. Our results of prospective disease surveillance in Panama indicate that 11.9% of suspected dengue cases are VEEV ID subtype infections and provide evidence for overlapping alphavirus circulation with other endemic arboviral infections such as dengue. Moreover, our findings support previous evidence suggesting that VEEV complex infections represent 10 % of the dengue burden in Latin American endemic countries(2). VEEV complex infections are clinically undistinguished from endemic infections such as dengue (2), thus the VEEV complex burden may be underestimated in dengue endemic regions.

VEEV ID subtype RNA was detected in three mosquito pools from the *Cx. (Mel*.*) vomerifer* species trapped during the 2015 outbreak in El Real de Santamaria, Panama. Mosquitoes, from the *Cx. (Mel*.*) vomerifer* have been previously incriminated as a vector for VEEV ID subtype in Panama (2). Two of these mosquito pools yielded viral isolates. None of the original pan-alphavirus convectional RT-PCRs were able to detect viral RNA in the mosquito pools. This suggests that our newly designed rRT-PCR has an increased sensitivity to detect VEEV complex RNA in mosquito vectors. No MADV nor EEEV infections were detected in mosquitoes by either pan-alpha, rRT-PCR or isolation methods. The same pattern of lack of MADV isolation or detection in mosquitoes was observed during previous outbreak investigations undertaken by our group in Panama (37, 38). Interestingly, low MADV frequency of isolation was also observed during extensive mosquito investigations undertaken during the 1940-60s in Panama by the Gorgas Memorial Laboratory; in these occasions MADV was only isolated twice from *Cx. (Mel*.*) taeniopus* in 1964 and 1973, respectively (39, 40). The low frequency of MADV detection or isolation in Panama contrast with features observed in the endemic region of Iquitos, Peru, where active circulation of MADV in the enzootic vector *Culex (Mel*.*) pedroi* is frequently detected(9, 41). Taken together, the early reports along with recent evidence suggest that mosquito vectors in Panama may be infected with MADV at a lower frequency when compared with VEEV ID subtype and even MADV in other endemic regions. The reasons for this variation in MADV and VEEV ID subtype frequency of isolation or detection from mosquitoes collected in Panama remain unclear and may include variation in vector competence or viral heterologous competition or even enhanced VEEV ID subtype transmission by insect specific virus as has been shown with dengue and Zika viruses (42).

Only a small number of human and mosquito samples were available to validate the assays. However, prospective disease surveillance allowed us to obtain additional samples to further validate our new assays. While previously developed methods relied on the validation of assays using plasmids, viral isolates, or a few human sera samples (13–21), our assay validation with human sera, mosquitos, and post-mortem tissue samples is more realistic and typical in reference laboratories. Moreover, our approach failed to detect two samples that had previously tested positive back in 2015 using standard alphavirus generic primers (19). Vina-Rodriguez et al., reported an rRT-PCR that was designed from an alignment of 33 VEEV sequences (21). Development of this assay did not include other species that comprise the VEEV complex, and clinical samples were not available for evaluation (21). Our assays were designed using a greater number of complete genome sequences with confirmation *in silico* that the primers and probes match contemporary alignments. Interestingly, a second round of the generic alphavirus RT-PCR undertaken in 2017 with the same primers have also failed to reamplify the former positives. Viral RNA degradation over time is a likely explanation of these false negative samples(43). Untargeted metagenomic sequencing of complete and near-complete genome sequences confirmed that the VEEV ID subtype was detected using the VEEV complex primers; this subtype has been detected in central and eastern Panama regions (7). Further VEEV ID subtype evolutionary analyses are currently being undertaken to investigate host or vector adaptations. Taken together, these data demonstrate the potential of molecular and genomic approaches to improve the detection of VEE complex, MADV and EEEV acute infections, even in long-time storage samples.

Additional prospective testing is warranted to fully characterize the assays’ clinical and surveillance performance. An additional limitation of this study is that the design requires two separate assays for three viruses as the optimal design targets for the VEEV complex and MADV overlap in a highly conserved region. However, the two rRT-PCRs can be performed together on a single run, however, which improves lab workflow, and the VEEV complex assay can be multiplexed with rRT-PCRs for other neurotropic arboviruses such as West Nile and St. Louis encephalitis viruses without a loss in performance (manuscript in preparation).

In conclusion, we describe new sensitive and specific VEEV complex, MADV and EEEV rRT-PCRs that provide significant benefits over available molecular methods, allowing us to detect VEEV-MADV co-infections and VEEV human infection in samples that were negative with other techniques. Early acute samples that were negative with other techniques. Early acute samples (≤ 3 days since onset of symptoms), with coincidental VEEV specific antibodies and VEEV viral RNA suggest that VEEV re-infections are possible. In addition, our newly developed method allowed us to detect VEEV active viral circulation in mosquitoes collected during an alphavirus outbreak response. The implementation of then assays in regions of endemicity may improve the identification and characterization of these neurotropic alphaviruses.

## Data Availability

All data produced in the present work are contained in the manuscript

## Acknowledgements

We thank Xacdiel Rodriguez, Yelissa Rios, Yaneth Pittí, Oriel Lezcano and Eddier Rivera and Mileika Santos for technical support with sample processing and mosquito classification and Alberto Cumbrera for the map construction. We also thank Leyda Abrego for providing reagents for the rRT-PCR and Milena Gomez, Thais M. Coletti, Esmenia Rocha, Geovana Maria Pererira, Erika R. Manuli for technical support with metagenomic sequencing

## References

1. Navarro JC, Carrera JP, Liria J, Auguste AJ, Weaver SC. 2017. Alphaviruses in Latin America and the introduction of chikungunya virusHuman Virology in Latin America: From Biology to Control.

2. Aguilar P v., Estrada-Franco JG, Navarro-Lopez R, Ferro C, Haddow AD, Weaver SC. 2011. Endemic Venezuelan equine encephalitis in the Americas: Hidden under the dengue umbrella. Future Virol https://doi.org/10.2217/fvl.11.50.

3. Arrigo NC, Adams AP, Weaver SC. 2010. Evolutionary Patterns of Eastern Equine Encephalitis Virus in North versus South America Suggest Ecological Differences and Taxonomic Revision. J Virol https://doi.org/10.1128/jvi.01586-09.

4. Forshey BM, Guevara C, Laguna-Torres VA, Cespedes M, Vargas J, Gianella A, Vallejo E, Madrid C, Aguayo N, Gotuzzo E, Suarez V, Morales AM, Beingolea L, Reyes N, Perez J, Negrete M, Rocha C, Morrison AC, Russell KL, Blair PJ, Olson JG, Kochel TJ. 2010. Arboviral etiologies of acute febrile illnesses in western south America, 2000-2007. PLoS Negl Trop Dis 4.

5. Carrera J-P, Forrester N, Wang E, Vittor AY, Haddow AD, López-Vergès S, Abadía I, Castaño E, Sosa N, Báez C, Estripeaut D, Díaz Y, Beltrán D, Cisneros J, Cedeño HG, Travassos da Rosa AP, Hernandez H, Martínez-Torres AO, Tesh RB, Weaver SC. 2013. Eastern Equine Encephalitis in Latin America. New England Journal of Medicine https://doi.org/10.1056/nejmoa1212628.

6. Carrera JP, Pittí Y, Molares-Martínez JC, Casal E, Pereyra-Elias R, Saenz L, Guerrero I, Galué J, Rodriguez-Alvarez F, Jackman C, Pascale JM, Armien B, Weaver SC, Donnelly CA, Vittor AY. 2020. Clinical and serological findings of madariaga and Venezuelan equine encephalitis viral infections: A follow-up study 5 years after an outbreak in Panama. Open Forum Infect Dis https://doi.org/10.1093/ofid/ofaa359.

7. Quiroz E, Aguilar P v., Cisneros J, Tesh RB, Weaver SC. 2009. Venezuelan equine encephalitis in Panama: Fatal endemic disease and genetic diversity of etiologic viral strains. PLoS Negl Trop Dis https://doi.org/10.1371/journal.pntd.0000472.

8. Corniou B, Ardoin P, Bartholomew C, Ince W, Massiah V. 1972. First isolation of a South American strain of Eastern Equine virus from a case of encephalitis in Trinidad. Trop Geogr Med 24.

9. Aguilar P v., Robich RM, Turell MJ, O’Guinn ML, Klein TA, Huaman A, Guevara C, Rios Z, Tesh RB, Watts DM, Olson J, Weaver SC. 2007. Endemic eastern equine encephalitis in the Amazon region of Peru. American Journal of Tropical Medicine and Hygiene https://doi.org/10.4269/ajtmh.2007.76.293.

10. Vittor AY, Armien B, Gonzalez P, Carrera J-P, Dominguez C, Valderrama A, Glass GE, Beltran D, Cisneros J, Wang E, Castillo A, Moreno B, Weaver SC. 2016. Epidemiology of Emergent Madariaga Encephalitis in a Region with Endemic Venezuelan Equine Encephalitis: Initial Host Studies and Human Cross-Sectional Study in Darien, Panama. PLoS Negl Trop Dis 10.

11. Gil LHVG, Magalhaes T, Santos BSAS, Oliveira L v., Oliveira-Filho EF, Cunha JLR, Fraiha ALS, Rocha BMM, Longo BC, Ecco R, Faria GC, Furtini R, Drumond SRM, Maranhão RPA, Lobato ZIP, Guedes MIMC, Teixeira RBC, Costa EA. 2021. Active circulation of madariaga virus, a member of the eastern equine encephalitis virus complex, in northeast brazil. Pathogens 10.

12. Lednicky JA, White SK, Mavian CN, el Badry MA, Telisma T, Salemi M, OKech BA, Beau De Rochars VM, Morris JG. 2019. Emergence of Madariaga virus as a cause of acute febrile illness in children, Haiti, 2015-2016. PLoS Negl Trop Dis https://doi.org/10.1371/journal.pntd.0006972.

13. Pfeffer M, Proebster B, Kinney RM, Kaaden OR. 1997. Genus-specific detection of alphaviruses by a semi-nested reverse transcription-polymerase chain reaction. American Journal of Tropical Medicine and Hygiene 57.

14. Brightwell G, Brown JM, Coates DM. 1998. Genetic targets for the detection and identification of Venezuelan equine encephalitis viruses. Arch Virol 143.

15. Romeiro MF, de Souza WM, Tolardo AL, Vieira LC, Henriques DA, de Araujo J, Siqueira CEH, Colombo TE, Aquino VH, da Fonseca BAL, de Morais Bronzoni RV, Nogueira ML, Durigon EL, Figueiredo LTM. 2016. A real-time RT-PCR for rapid detection and quantification of mosquito-borne alphaviruses. Arch Virol 161.

16. Linssen B, Kinney RM, Aguilar P, Russell KL, Watts DM, Kaaden OR, Pfeffer M. 2000. Development of reverse transcription-PCR assays specific for detection of equine encephalitis viruses. J Clin Microbiol 38.

17. Wang E, Paessler S, Aguilar P v., Carrara AS, Ni K, Greene IP, Weaver SC. 2006. Reverse transcription-PCR-enzyme-linked immunosorbent assay for rapid detection and differentiation of alphavirus infections. J Clin Microbiol 44.

18. Pisano MB, Seco MPS, Ré VE, Farías AA, Contigiani MS, Tenorio A. 2012. Specific detection of all members of the Venezuelan Equine Encephalitis complex: Development of a RT-Nested PCR. J Virol Methods 186.

19. Sánchez-Seco MP, Rosario D, Quiroz E, Guzmán G, Tenorio A. 2001. A generic nested-RT-PCR followed by sequencing for detection and identification of members of the alphavirus genus. J Virol Methods https://doi.org/10.1016/S0166-0934(01)00306-8.

20. Bronzoni RVM, Moreli ML, Cruz ACR, Figueiredo LTM. 2004. Multiplex nested PCR for Brazilian Alphavirus diagnosis. Trans R Soc Trop Med Hyg 98.

21. Vina-Rodriguez A, Eiden M, Keller M, Hinrichs W, Groschup MH. 2016. A quantitative real-time RT-PCR assay for the detection of venezuelan equine encephalitis virus utilizing a universal alphavirus control RNA. Biomed Res Int 2016.

22. Benson DA, Cavanaugh M, Clark K, Karsch-Mizrachi I, Ostell J, Pruitt KD, Sayers EW. 2018. GenBank. Nucleic Acids Res https://doi.org/10.1093/nar/gkx1094.

23. Rojas A, Cardozo F, Cantero C, Stittleburg V, López S, Bernal C, Gimenez Acosta FE, Mendoza L, Pinsky BA, de Guillén IA, Páez M, Waggoner J. 2019. Characterization of dengue cases among patients with an acute illness, Central Department, Paraguay. PeerJ 2019.

24. Claudino-Sales V. 2019. Darien National Park, PanamaCoastal Research Library.

25. Santiago GA, Vázquez J, Courtney S, Matías KY, Andersen LE, Colón C, Butler AE, Roulo R, Bowzard J, Villanueva JM, Muñoz-Jordan JL. 2018. Performance of the Trioplex real-time RT-PCR assay for detection of Zika, dengue, and chikungunya viruses. Nat Commun 9.

26. Sallum MAM, Forattini OP. 1996. Revision of the spissipes section of culex (melanoconion) (diptera: Culicidae). J Am Mosq Control Assoc.

27. Johnson BW, Kosoy O, Wang E, Delorey M, Russell B, Bowen RA, Weaver SC. 2011. Use of Sindbis/eastern equine encephalitis chimeric viruses in plaque reduction neutralization tests for arboviral disease diagnostics. Clinical and Vaccine Immunology 18.

28. Claro IM, Ramundo MS, Coletti TM, da Silva CAM, Valenca IN, Candido DS, Sales FCS, Manuli ER, de Jesus JG, de Paula A, Felix AC, Andrade P dos S, Pinho MC, Souza WM, Amorim MR, Proenca-Modena JL, Kallas EG, Levi JE, Faria NR, Sabino EC, Loman NJ, Quick J. 2021. Rapid viral metagenomics using SMART-9N amplification and nanopore sequencing. Wellcome Open Res 6.

29. Li H. 2018. Minimap2: Pairwise alignment for nucleotide sequences. Bioinformatics 34.

30. Li H, Handsaker B, Wysoker A, Fennell T, Ruan J, Homer N, Marth G, Abecasis G, Durbin R. 2009. The Sequence Alignment/Map format and SAMtools. Bioinformatics 25.

31. Oxford Nanopore. 2022. GitHub Create statistic summary of an Oxford Nanopore read dataset.

32. Milne I, Stephen G, Bayer M, Cock PJA, Pritchard L, Cardle L, Shawand PD, Marshall D. 2013. Using tablet for visual exploration of second-generation sequencing data. Brief Bioinform 14.

33. Katoh K, Standley DM. 2013. MAFFT multiple sequence alignment software version 7: Improvements in performance and usability. Mol Biol Evol 30.

34. Minh BQ, Schmidt HA, Chernomor O, Schrempf D, Woodhams MD, von Haeseler A, Lanfear R, Teeling E. 2020. IQ-TREE 2: New Models and Efficient Methods for Phylogenetic Inference in the Genomic Era. Mol Biol Evol 37.

35. Luciani K, Abadía I, Martínez-Torres AO, Cisneros J, Guerra I, García M, Estripeaut D, Carrera JP. 2015. Case report: Madariaga virus infection associated with a case of acute disseminated encephalomyelitis. American Journal of Tropical Medicine and Hygiene 92.

36. Torres-ruesta A, Chee RSL, Ng LFP. 2021. Insights into antibody-mediated alphavirus immunity and vaccine development landscape. Microorganisms https://doi.org/10.3390/microorganisms9050899.

37. Torres R, Samudio R, Carrera JP, Young J, Maârquez R, Hurtado L, Weaver S, Chaves LF, Tesh R, Caâceres L. 2017. Enzootic mosquito vector species at equine encephalitis transmission foci in the República de Panama. PLoS One https://doi.org/10.1371/journal.pone.0185491.

38. Carrera J-P, Cucunuba ZM, Neira K, Lambert B, Pitti Y, Liscano J, Garzon JL, Beltran D, Collado-Mariscal L, Saenz L, Sosa N, Rodriguez-Guzman LD, Gonzalez P, Lezcano AGAG, Pereyra-Elias R, Valderrama A, Weaver SC, Vittor AY, Armien B, Pascale J-M, Donnelly CA. 2020. Endemic and epidemic human alphavirus infections in eastern Panama: An analysis of population-based cross-sectional surveys. American Journal of Tropical Medicine and Hygiene 10:901462.

39. Dietz WH, Galindo P, Johnson KM. 1980. Eastern equine encephalomyelitis in Panama: The epidemiology of the 1973 epizootic. American Journal of Tropical Medicine and Hygiene https://doi.org/10.4269/ajtmh.1980.29.133.

40. Srihongse S, Galindo P. 1967. The isolation of eastern equine encephalitis virus from Culex (Melanoconion) taeniopus Dyar and Knab in Panama. Mosquito News 27:74–76.

41. Turell MJ, O’Guinn ML, Jones JW, Sardelis MR, Dohm DJ, Watts DM, Fernandez R, Travassos Da Rosa A, Guzman H, Tesh R, Rossi CA, Ludwig G v., Mangiafico JA, Kondig J, Wasieloski LP, Pecor J, Zyzak M, Schoeler G, Mores CN, Calampa C, Lee JS, Klein TA, 2006. Isolation of Viruses from Mosquitoes (Diptera: Culicidae) Collected in the Amazon Basin Region of Peru. J Med Entomol 42:891–898.

42. Olmo RP, Todjro YMH, Aguiar ERGR, de Almeida JPP, Ferreira F v., Armache JN, de Faria IJS, Ferreira AGA, Amadou SCG, Silva ATS, de Souza KPR, Vilela APP, Babarit A, Tan CH, Diallo M, Gaye A, Paupy C, Obame-Nkoghe J, Visser TM, Koenraadt CJM, Wongsokarijo MA, Cruz ALC, Prieto MT, Parra MCP, Nogueira ML, Avelino-Silva V, Mota RN, Borges MAZ, Drumond BP, Kroon EG, Recker M, Sedda L, Marois E, Imler JL, Marques JT. 2023. Mosquito vector competence for dengue is modulated by insect-specific viruses. Nat Microbiol 8:135–149.

43. Relova D, Rios L, Acevedo AM, Coronado L, Perera CL, Pérez LJ. 2018. Impact of RNA degradation on viral diagnosis: An understated but essential step for the successful establishment of a diagnosis network. Vet Sci 5.

